# Maternal Serum Concentrations of Selenium, Copper and Zinc during Pregnancy are associated with Risk of Spontaneous Preterm Birth: A case-control study from Malawi

**DOI:** 10.1101/2020.04.14.20064717

**Authors:** Grace Chiudzu, Augustine T. Choko, Alfred Maluwa, Sandra Huber, Jon Odland

## Abstract

Preterm birth is delivery before 37 completed weeks. A study was conducted to evaluate the association of maternal serum concentrations of selenium, copper, and zinc, and preterm birth. There were 181 women in this nested case-control study, 90/181 (49.7%) term and 91/181 (50.3%) preterm pregnant women. The overall mean serum concentration of selenium was 77.0; SD 19.4µg/L, copper was 2.50; SD 0.52 mg/L and zinc was 0.77; SD 0.20 mg/L with reference values of 47-142µg/L, 0.76-1.59mg/L and 0.59-1.11 mg/L, respectively. For preterm birth, mean serum concentrations for selenium was 79.7; SD 21.6µg/L, copper was 2.61; SD 0.57 mg/L, and zinc was 0.81; SD 0.20 mg/L compared to that of term births: selenium (74.2; SD 16.5µg/L; p=0.058), copper (2.39; SD 0.43 mg/L; p = 0.004), and zinc (0.73; SD 0.19 mg/L; p = 0.006) respectively. In adjusted analysis, every unit increase in maternal selenium concentrations gave increased odds of being a case OR 1.01 (95% CI: 0.99; 1.03), p=0.234, copper OR 1.62 (95% CI: 0.80; 3.32); p = 0.184, zinc OR 6.88 (95% CI: 1.25; 43.67); p=0.032. Results show that there was no deficiency of selenium, and zinc; and high serum concentrations of copper in pregnancy. Preterm birth was associated with higher maternal serum concentrations of copper and zinc.

## Introduction

Preterm birth occurs before 37 completed weeks (259 days) after the first day of the last menstrual period preceding the pregnancy [1]. Globally, it is estimated that 14.9 million babies are born preterm every year. with an average preterm birth rate of 11.1% per year [2]. Variations are present in different regions, ranging from 5% in western countries to 18% in some Africa countries with, Malawi being reported at 18.1% [3]. Preterm birth has adverse effects and accounts for over 75% of perinatal mortality and more than 50% of morbidity with long term health consequences like intellectual disabilities, cerebral palsy, vision/hearing impairments; and complications of respiratory, gastrointestinal and renal systems due to immaturity of multiple organ systems [4]–[6]. In addition, studies have shown significant association between preterm birth and the risk of chronic degenerative diseases such as hypertension, coronary heart disease, and type 2 diabetes mellitus during adulthood[7].

Preterm birth is a condition with many pathological processes and no precise mechanism. A common pathway involving cellular apoptosis transmitting inflammatory signal, senescent placental cells and its changes in the placental membrane were hypothesised to stimulate parturition both term and preterm [8]. This common pathway is believed to be stimulated by the premature aging of the placenta caused by oxidative stress [9]. The placenta is the main source of reactive oxygen species (ROS) which induce cellular damage by acting on proteins and lipids [9]. Excessive production of ROS or impaired antioxidative defence results in a variety of pregnancy complications like preeclampsia and preterm birth[9]–[11]. Infections also up-regulate ROS, increasing the risk of preterm birth. The micronutrients of selenium (Se), copper (Cu) and zinc (Zn) act as cofactors of antioxidants enzymes that counterbalances the oxidative stress and regulate the inflammatory response [13], [14].

Micronutrients, though required in minute quantities, are necessary inorganic constituents of human health. Their concentration status could lead to adverse pregnancy outcomes. Selenium is integrated into proteins to make selenoproteins which includes glutathione peroxidase, thioredoxin reductases and selenoprotein-P [15]. Reduced levels of maternal selenium concentrations have been shown to be associated with recurrent early pregnancy loss and preeclampsia[16]–[20]. Copper is an essential cofactor for numerous enzymes involved in various biological processes. The effect of maternal copper concentration is not well understood with some studies suggesting increased preterm birth with copper deficiency [21]–[24] and others reporting no association [25], [26]. Zinc is a crucial component of many metalloenzymes participating in protein and carbohydrate metabolism, nucleic acid synthesis and antioxidants functions through Cu/Zn superoxide dismutase [27]. It also assists with fetal brain development during pregnancy, as well as being an aid to the mother during labour [28]. Changes in zinc homeostasis have been associated with various effects on pregnancy including prolonged labour, fetal growth restriction, fetal death, preeclampsia and preterm birth[29]–[33].

Haematological concentrations of micronutrients are influenced by diet, lifestyle and environmental conditions. Whether maternal concentrations of selenium, copper and zinc during pregnancy have an effect on preterm birth in sub-Saharan Africa needs to be further determined. Subsequently, this study was conducted to evaluate the status of selenium, copper and zinc in pregnancy and whether their concentrations are different between term and preterm pregnancies in a Malawian population. The knowledge gained on the exact role these micronutrients play in pregnancy would assist in improving nutrition in pregnancy to reduce the incidence of pregnancy complications specifically preterm birth.

The introduction should be succinct, with no subheadings. Limited figures may be included only if they are truly introductory, and contain no new results.

## Materials and Methods

This nested case control study was conducted between June 2016 and March 2017 at Kamuzu Central Hospital and Bwaila Hospital, tertiary and general hospitals respectively The two hospitals are public and government funded and both located within the city of Lilongwe, the capital of Malawi in the central region. Spontaneous preterm birth patients within the district can be managed at either hospital. The early preterm (gestation of <34 weeks) are referred to Kamuzu Central Hospital because of the availability of neonatal services while the late preterm (>34 - <37weeks) are managed at Bwaila Hospital. Therefore the two hospitals were chosen in order to capture both early and late spontaneous preterm birth. Our main outcome was to determine the concentrations of selenium, copper and zinc in term and spontaneous preterm births. The secondary outcome was to determine the association of spontaneous preterm birth and maternal serum concentrations of selenium, copper and zinc.

The target populations were all pregnant women presenting with spontaneous preterm birth at these sites. Gestational age was based on prior dating ultrasound. Women coming without ultrasound dated gestation, ultrasound was done at presentation prior to recruitment and Ballard score was done after delivery to exclude small for gestation age babies[34]. The cases were defined as preterm pregnancies of gestation 26 - <37 weeks while controls were term pregnancies of gestation 37 - ≤41 completed weeks. The inclusion criteria were the following: singleton pregnancy, presenting in spontaneous labour with intact membranes and willing to participate in the study. Exclusion criteria included women having medically indicated termination of pregnancy, multiple gestations, and pregnancy complications like abruption placentae, preeclampsia, and clinical evidence of chorioamnionitis as these would cause preterm birth. Recruitment was done by a trained midwife while on duty, day or night including the weekends. Systemic random sampling was used with numbers allocated to all eligible preterm cases and all with odd numbers recruited in the study. For controls they were recruited as they present in the labour ward matched to the living location.

Soon after delivery process, the trained midwife proceeded to withdraw 5 mL of venous blood using a regular vacutainer technique into a royal blue top vacutainer tube (lot No. BD368380). The blood samples were immediately transported to Baylor project laboratory within the hospital premises where centrifuging at 3000 g for 10 minutes was done. Serum was transferred into a screw-top vial and stored in the freezer at −80 °C. Metal free vessels were used throughout the process. When the required number of specimens was reached, the serum samples were shipped in the frozen state to the the University Hospital of North Norway (UNN), Tromsø, Norway for chemical analysis.

A structured questionnaire was administered in vernacular language (Chichewa) - a local language which study participants were conversant with, by the trained midwife. Quantitative data were captured on demographic characteristics of the participants: - age, gravidity, marital status, educational level, and previous history of preterm birth. Information on antenatal care package including; gestation age, serology test results for human immunodeficiency virus (HIV), and syphilis were collected from the patient’s health passport. Self-reported information on herbal medicine ingestion defined by World Health Organisation (WHO) as “any medicinal product based on herbs, herbal materials, herbal preparations and finished products that contain as active ingredients parts of plants, other than plant materials, or combinations thereof” and bedside laboratory test of haemoglobin using Hemocue were documented in the questionnaire.

The chemical analyses were conducted at the Norwegian laboratory in Tromsø, using inductively coupled plasma mass spectrometry (ICP-MS) for analysis of the trace elements selenium (Se), copper (Cu), and zinc (Zn). For sample preparation, an automated liquid handler Tecan Freedom Evo 200 (Männedorf, Switzerland) equipped with an 8- channel liquid handler arm (LiHa) for conductive disposable tips, a robotic manipulator arm (RoMa) for transport of microtiterplates and a shaker (Bioshake, Quantifol instruments GmBH, Jena, Germany) were used. 100 µL serum was diluted with a solution consisting of Milli-Q water (Millipore/ Merck KGaA, Darmstadt, Germany) with 0.08% v/v Triton X-100 (Sigma/ Merck KGaA, Darmstadt, Germany), 10% v/v ammonium (Honeywell Fluka, Bucharest,Romania) and isopropanol (Honeywell Fluka, Bucharest, Romania) and 0.25 µg/L gold (Au; Inorganic Ventures, Christiansburg, VA, USA) and mixed on the bioshaker for 2 minutes. The instrumental analysis was performed on a NEXION 300D ICP-MS system (Perkin Elmer, Waltham, Massachusetts, USA) equipped with an ESI-Fast SC2DX auto sampler. An internal standard solution was introduced online via a T-piece containing 20 µg/L rhodium (Rh^103^; Inorganic Ventures, Christiansburg, VA, USA). For the MS analysis the kinetic-energy-discrimination (KED) mode was applied. Measurements were conducted in triplicates. All concentrations were obtained by the internal standard method together with a blank subtraction using the NexION software (version 1.5). The trace element concentrations within the samples were determined based on a 3-point external matrix matched calibration curve using ClinCal serum calibration material from Recipe (Recipe, Munich, Germany), which was diluted together with each batch of 32 samples by 1:100, 1:40 and 1:20 respectively. For quality assurance and quality control two sets of ClinCheck control material L-1 and L-2 from Recipe (Recipe, Munich, Germany), and Seronorm L-1 and L-2 (Sero, Billingstad, Norway), as well as 4 Milli-Q water blanks were run together with each batch of samples. Diluent blanks for control of the background and instrumental carry over were also included. Additionally, the Laboratory for Analysis of Environmental Pollutants, UNN, Norway, participates successfully in the international quality control programme: Quebec Multielement External Quality Assessment Scheme (QMEQAS) organized by the Centre de toxicologie du Quebec, Quebec, Canada.

The study was reviewed and approved by College of Medicine Research Committee (COMREC) P.05/15/1738. In addition, the Directors of Kamuzu Central and Bwaila Hospitals approved the study to take place at their respective hospitals. Written informed consent was obtained from all study participants.

### 2.1 Data management and analysis

Quantitative data from the questionnaires and laboratory reports were entered into IBM SPSS version 14.0 software by a data entry clerk and verified by the principal investigator. Data cleaning was performed involving a range of consistency checks, and all outliers were rectified

Data analysis was done using R (R Core team, 2015). A binary variable was generated for the primary outcome based on gestation: 26 - <37 weeks (preterm birth as cases) coded as 1 and ≥37 - 41 weeks (term birth as controls) as 0. One-way frequencies were computed for categorical variables while mean and standard deviation (SD) or median and inter quartile range [IQR] were computed for continuous variables that were normally distributed or skewed, respectively. Log transformation was done where necessary for positively skewed continuous variables.

Two-way associations between independent variables and the outcome variable of preterm birth were investigated using the Pearson’s correlation for continuous independent variables and the Chi squared test for categorical variables. P-values from all tests of less than 0.05 were considered statistically significant. Logistic regression models were fitted, addressing each covariate with the outcome variable. Variables were included in the multiple logistic regression model based on the best fit regression model from stepwise regression analysis using the R software based on the observed association in the unadjusted analysis at the p<0.05 test level of significance. Odds ratios and associated 95% confidence intervals (CI) were computed from the logistic regression models.

The materials and methods section should contain sufficient detail so that all procedures can be repeated. It may be divided into headed subsections if several methods are described.

## Results and Discussion

There were 181 mothers with overall mean maternal age of 25.2 years (SD: 6.1); and 181 babies participating in the present study (Table 1). The majority were married or cohabitating (173/181, 96%) and presented in the first or second pregnancy (108/181, 60%). Fifty per cent had attained some primary education. Overall 9% (16/181) gave a history of delivery of previous preterm birth while 17.7% (32/181) reported having used herbal medications in the current pregnancy. Human immunodeficiency virus and syphilis were present in (27/181, 15%) and (1/181, 0.6%) respectively.

**Table 1:** Characteristics of study participants for term versus preterm births

| Variable | characteristic | Total<br>n=181 | Term births<br>n=90 | Preterm<br>births<br>n=91 | p-value |
| --- | --- | --- | --- | --- | --- |
| Maternal age (years). | Mean (SD) | 25.2 (6.1) | 26.32 (5.92) | 24.01 (6.19) | 0.011* |
| Gestation (weeks) | Mean (SD) | 35.6 (1.8) | 38.00 (1.04) | 33.36 (2.55) | <0.0001* |
| Birth weight (kg) | Mean (SD) | 2.6 (0.4) | 3.14 (0.38) | 2.12 (0.48) | <0.0001* |
| Married/cohabitating | N (%) | 173 (96) | 87 ( 96.7) | 86 (94.5) | 0.766 |
| Gravidity | N (%) |  |  |  | 0.048* |
| • 1-2 |  | 108 (60) | 46 ( 51.1) | 62 (68.1) |  |
| • 3-5 |  | 61 (34) | 38 ( 42.2) | 23 (25.3) |  |
| • $\geq 6$ | | 12 (6.6) | 6 (6.7) | 6 (6.6) | |
| Preterm baby before | N (%) | 16 (9) | 4 (4.4) | 12 (13.2) | 0.38 |
| Education Level | N (%) |  |  |  | 0.31 |
| • ≥College | | 11 (6.1) | 5 ( 5.6) | 6 (6.6) | |
| • Secondary |  | 70 (38.7) | 41 ( 45.6) | 29 (31.9) |  |
| • Primary |  | 91 (50.3) | 40 ( 44.4) | 51 (56.0) |  |
| • none |  | 9 (5) | 4 ( 4.4) | 5 (5.5) |  |
| Herbal medication use | N (%) | 32 (17.7) | 10 ( 11.1) | 22 (24.2) | 0.035* |
| HIV positive | N (%) | 27 (15) | 14 ( 15.6) | 13 (14.3) | 0.975 |
| Syphilis positive | N (%) | 1 (0.6) | 0 ( 0.0) | 1 (1.1) | 0.018* |

SD: standard deviation, N: number, kg: kilograms
t-test used for continuous variables, Chi-square or Fisher's Exact test used for categorical variables

### 3.1 Two-way associations with being preterm birth

There were 90/181 (49.7%) term and 91/181 (50.3%) spontaneous preterm births in the present study (Table 1). Women delivering at term were older, with a mean age of 26.3 years (SD: 5.9) compared to 24.0 years (SD: 6.2), p = 0.011 for women who had preterm births. Women who had spontaneous preterm births were more likely to have low gravidity than those who had a term birth (p = 0.048). Use of herbal medications was significantly higher among women with spontaneous preterm birth (22/91, 24.2%) than among those with term birth (10/90, 11.1%), p = 0.035. Syphilis was significantly associated with being a preterm birth p = 048.

Two way analyses were also investigated for a number of trace elements (Table 2). The overall mean concentration values of selenium (77.0µg/L; SD 19.4µg/L) and zinc (0.77mg/L; SD 0.20 mg/L) were within the normal laboratory reference values (47-142µg/L) and (0.59 - 1.11 mg/L) respectively. The overall mean concentration value of copper (2.50 mg/L; SD 0.52 mg/L) was above normal laboratory reference value (0.76 - 1.59 mg/L). All concentrations of trace elements were higher among cases compared to controls. Selenium concentrations were marginally increased for women with spontaneous preterm birth (mean 79.7µg/L; SD: 21.6µg/L) compared to those with term births (mean 74.2µg/L; SD: 16.5µg/L), p = 0.058. Copper and zinc concentrations were significantly higher for women with spontaneous preterm births than those with term births with mean of 2.61 mg/L (SD: 0.57 mg/L) vs. 2.39 mg/L (SD: 0.43 mg/L), p = 0.004 for copper and a mean of 0.81 mg/L (SD: 0.20 mg/L) vs. 0.73 mg/L (SD: 0.19 mg/L), p = 0.006 for zinc

**Table 2:** Two way analysis for micronutrient concentrations

| Serum concentration | characteristic | Lab Ref Values[35] | Total n=181 | Term births n=90 (mg/L) | Preterm births n=91 | p-value |
| --- | --- | --- | --- | --- | --- | --- |
| Selenium ($\mu\text{g/L}$) | Mean (SD) | 47-142 | 77.0 (19.4) | 74.2(16.5) | 79.7(21.6) | 0.058 |
| Copper (mg/L) | Mean (SD) | 0.76-1.59 | 2.50 (0.52) | 2.39 (0.43) | 2.61 (0.57) | 0.004 |
| Zinc (mg/L) | Mean (SD) | 0.59-1.11 | 0.77 (0.20) | 0.73 (0.19) | 0.81 (0.20) | 0.006 |

SD: standard deviation
t-test used for continuous variables

In adjusted analysis, similar results to those for unadjusted analysis were observed for the metals (Table 3). There was no association between a unit increase in selenium concentration and increased odds of being a spontaneous preterm birth, OR 1.01(95% CI: 0.99; 1.03, p = 0.234). Although there was an association between a unit increase in copper and increased odds of being a spontaneous preterm birth in unadjusted analysis (OR 2.38, CI: 1.31; 4.54, p = 006), this association was no longer significant in adjusted analysis, OR 1.62 (95% CI: 0.80; 3.32, p = 0.184). However, there was an association between a unit increase in zinc and increased risk for being a preterm birth in both unadjusted, OR 8.56 (95% CI: 1.83; 46.10, p = 009) and adjusted analysis, OR6.88 (95% CI: 1.25; 43.67, p = 0.032). Women who had taken herbal medication had increased odds of having a spontaneous preterm birth compared to those who did not take the herbal medication, however this was not significant, OR2.44 (95% CI:0.97, 6.52, p=0.064).

**Table 3:** Unadjusted and adjusted estimates for being preterm (complete case analysis)

| Variable | Characteristic | Unadjusted |  |  |  | Adjusted |  |  |  |
| --- | --- | --- | --- | --- | --- | --- | --- | --- | --- |
|  |  | OR | 95% CI |  | p-value | OR | 95% CI |  | p-value |
| Maternal age | Yearly increase | 0.94 | 0.89 | 0.99 | 0.012 | 0.99 | 0.92 | 1.07 | 0.862 |
| Herbal medication | No |  |  |  |  |  |  |  |  |
|  | Yes | 2.55 | 1.16 | 5.98 | 0.024 | 2.44 | 0.97 | 6.52 | 0.064 |
| Unit increase in Selenium | Unit | 1.02 | 1.00 | 1.03 | 0.063 | 1.01 | 0.99 | 1.03 | 0.234 |
| Unit increase in Copper | Unit | 2.38 | 1.31 | 4.54 | 0.006 | 1.62 | 0.80 | 3.32 | 0.184 |
| Unit increase in Zinc | Unit | 8.56 | 1.83 | 46.10 | 0.009 | 6.88 | 1.25 | 43.67 | 0.032 |
| Gravidity | 1-2 | ref |  |  |  |  |  |  |  |
|  | 2-5 | 0.45 | 0.23 | 0.85 | 0.015 | 0.68 | 0.29 | 1.61 | 0.381 |
|  | ≥6 | 0.74 | 0.22 | 2.51 | 0.624 | 1.14 | 0.21 | 6.12 | 0.874 |

OR: odds ratio; CI: confidence interval; ref: refer
Simple and multivariable logistic regression used.

The current study found that the overall serum concentrations for selenium and zinc were within normal reference values while that of copper was above normal reference values. Concentrations of selenium, copper and zinc were higher in women who had spontaneous preterm birth compared to those who delivered at term, with copper and zinc concentrations being significant. After adjusting for other variables increase in zinc concentration was associated with increased risk of spontaneous preterm birth.

Very little information was found in the literature on the maternal serum concentrations of the trace elements selenium, copper and zinc in pregnancy and their associations with spontaneous preterm birth in the African region. Comparing with researches from other regions some studies have found maternal concentrations of these trace elements to be associated with preterm birth, while others have reported no associations [21], [25], [36]– [40]. In this study, higher selenium concentrations were observed in maternal serum of preterm birth, but this was not significant. This finding is similar to the study by Irwinda et al. done in Indonesia, which recruited 51 women and found no difference in maternal selenium serum concentrations between term and preterm births[36]. Contrary to this study findings, a study in Lagos, Nigeria reported selenium deficiency of 20.4% at 14-26 weeks gestation and an 8-fold increase in risk of preterm delivery in an HIV positive population[37]. This contrast may be due to the differences in the study population and methodology with the Nigeria study being predominantly HIV positive population and samples taken during the second trimester. Likewise, in this study, women with preterm birth showed significantly higher maternal serum concentrations of copper but this was not associated with increased risk of being a preterm birth. This finding is consistent with findings of other studies reported in the literature[21], [25], [38]. Hao et al. collected plasma and serum at first prenatal visit between 4-22 gestation weeks and found that the overall medium maternal serum copper concentrations were significantly higher for preterm birth than term births in the Chinese population[39]. Contrary to the findings of other studies in the literature, this study found that maternal serum concentration of zinc were higher in women with preterm birth than those delivering at term and a unit increase of zinc had a 7-fold increase risk of having a preterm birth. A study conducted by Wang H et al. in a Chinese population found significant increased concentrations of zinc in women with term births compared to women with preterm births, p<0001[33]. However the study was prospective with samples collected during the first and second trimesters. Some studies have reported no association of maternal zinc concentrations and preterm birth[36], [41]. The difference may be due to study population and methodology used.

This study finding of increased maternal serum concentrations of selenium, copper and zinc in women with preterm birth is not surprising. As pregnancy progresses, the fetus take up selenium, copper, and zinc with maximum concentrations reached at the end of third trimester when the fetal liver is mature enough to store these trace elements[40], [42]–[44]. Spontaneous preterm birth reduces the pregnancy duration with resultant increased maternal serum concentrations and reduced fetal hepatic stores. Clinical research indicate that serum concentrations of selenium, copper, and zinc are higher in term infants than preterm infants[21]–[24], [44]–[46]. Consequently, with adequate nutrition, serum concentrations of selenium, copper and zinc would be expected to be higher in serum of women delivering preterm as there is reduced time for the mother to pass these trace elements to placenta and fetus. The lack of significant association of maternal serum concentrations of selenium and preterm birth may be due to the small number studied as a sample size of 600 was needed to see an effect in the present population.

The mechanism for the association of maternal serum concentration and increased risk of preterm birth could be explained due to the dual function of zinc, as an anti-oxidant as well as a pro-oxidant, and the cytotoxicity of copper. The placenta is the main source for ROS [9]. Nonetheless, the placenta is equipped with antioxidants inclusive of selenium-dependent enzymes of glutathione dismutase, thioredoxin reductases, selenoprotein-P, and copper/zinc superoxide dismutase which require the investigated trace elements of selenium, copper, and zinc [42]. However, the elevated free zinc and copper ion in women delivering preterm could have caused the generation of ROS. Free zinc ion damages mitochondria and NADPH oxidases to produce ROS[47]. Copper (I) ion is also capable of producing ROS by itself[48].

The finding of maternal serum copper concentration above the normal limit in both preterm and term deliveries also needs further investigation. Considering that excess copper ingestion has negative health effects on humans, the study recommends looking at the social habits, diet and water as exposure for excess copper ingestion.

Women who had taken herbal medication had almost 2.5 times increased risk of having spontaneous preterm birth compared to those who did not. It is possible that herbal medicine increased intake of zinc and copper ion in women which resulted in increased production of ROS. Further research is required to establish this link.

Some limitations should be considered when interpreting these study results. Firstly, the sample size was relatively small which reduced the power to 34% for detecting the effect. Recruitment was also only done when the research assistants were on duty and this may have affected the randomisation. Increased sample size and purposely recruited research assistants working in shifts to recruit day and night for the seven days of the week should be considered for future studies. Secondly, the study population diet information was not available as such it was difficult to ascertain the daily dietary intake of these trace elements. Thirdly, data on body mass index (BMI) was not captured in this study. This made the findings of this study not comparable with the findings by Broek et al. which noted the association of very low BMI with preterm birth[49]. Irrespective, the findings of no overall deficiencies of these trace elements in pregnancy and the fact that women delivering preterm babies had higher concentrations of the trace elements compared to those delivering at term, there is need to establish the causal link in preterm birth. The increased maternal serum copper concentrations in both term and preterm deliveries needs further investigations considering the harmful effects of excess copper.

## Conclusions

No selenium, copper, or zinc micronutrient deficiencies were found in the study population. Preterm births were associated with higher maternal serum concentrations of micronutrients copper and zinc which play a role in pro-oxidation, and anti-oxidation as well as anti-inflammatory mechanism. Copper maternal serum concentrations were above the upper normal limit in both term and preterm births groups. As such, further assessments are recommended to determine the excess source of copper. This study is a first assessment of these trace elements in mothers of term and preterm pregnancies in Malawian women. Further assessments in first and second trimesters with dietary information are recommended to establish local reference values as well as causal relationship.

## Data Availability

Data is available on request

## Data Availability

The SPSS data used to support the findings of this study are included within the supplementary information file

## Acknowledgments

We heartily acknowledge The Bill & Melinda Gates Foundation and Safe Motherhood Malawi through Dr Jennifer Tang for economic support of the study. The authors also like to thank Julie Sopie Kleppe Strømberg and Christina Ripman Hansen from the Department of Laboratory Medicine, University Hospital of North Norway, for assistance during sample preparation and the Helse Nord Malawi Fund for generously support of the analytical work.

## Supplementary Materials

CasesControlsmastercopytraceelements.sav

